# Multidisciplinary clinical assessment and interventions for childhood listening difficulty and auditory processing disorder: Relation between research findings and clinical practice

**DOI:** 10.1101/2024.06.12.24308837

**Authors:** David R. Moore, Li Lin, Ritu Bhalerao, Jody Caldwell-Kurtzman, Lisa L. Hunter

## Abstract

**Purpose:** Listening difficulty (LiD), often classified as auditory processing disorder (APD), has been studied in both research and clinic settings. The aim of this study was to examine the predictive relation between these two settings. In our “SICLiD” research study, children with normal audiometry, but caregiver-reported LiD, performed poorly on both listening and cognitive tests. Here we examined results of clinical assessments and interventions for these children in relation to research performance.

**Methods:** Study setting was a tertiary pediatric hospital. Electronic medical records were reviewed for 64 children aged 6-13 years recruited into a SICLiD LiD group based on a caregiver report (ECLiPS). The review focused on clinical assessments and interventions provided by Audiology, Occupational Therapy, Psychology (Developmental and Behavioral Pediatrics), and Speech-Language Pathology services, prior to study participation. Descriptive statistics on clinical encounters, identified conditions, and interventions were compared with quantitative, standardized performance on research tests. Z-scores were compared for participants with and without each clinical condition using univariate and logistic prediction analyses.

**Results:** Overall, 24 clinical categories related to LiD, including APD, were identified. Common conditions were attention (32%), language (28%), hearing (18%), anxiety (16%), and autism spectrum (6%) disorders. Performance on research tests varied significantly between providers, conditions, and interventions. Quantitative research data combined with caregiver reports provided reliable predictions of all clinical conditions except APD. Individual test significant correlations were scarce, but included the SCAN composite score, which predicted clinical language and attention difficulties, but not APD diagnoses.

**Conclusions:** The variety of disciplines, assessments, conditions and interventions revealed here supports previous studies showing that LiD is a multifaceted problem of neurodevelopment. Comparisons between clinical- and research-based assessments suggest a path that prioritizes caregiver reports and selected psychometric tests for screening and diagnostic purposes.

## Introduction

A primary goal of health research is to guide clinical practice. The aim of this study was to assess whether research findings from children with caregiver-reported listening difficulties (LiD), but without a hearing loss, predict clinical assessments and outcomes in the same group of children. To our knowledge, such an assessment has not previously been attempted.

LiD is an umbrella term that includes auditory processing disorder (APD), a deficit of auditory perception, operationally measured by performance on a variable series of tests of complex, supra-threshold auditory performance (Dillon and Cameron, 2021; Emanuel et al., 2011; Keith, 2009). APD is believed to originate in the brain, not the ear, since people diagnosed with APD typically do not have a clinical hearing loss or other otological disorders (ASHA, 2005; Audiology, 2010).

There have been many debates and discussions among health professionals and researchers about APD diagnosis (Dillon & Cameron, 2021; Illiadou & Kiese-Himmel, 2018; Moore, 2018) and treatment (Bellis et al., 2012; Fey et al., 2011) in school-aged children. APD (and LiD) overlap with developmental language disorders, attention deficit hyperactivity disorders, autism spectrum disorders, and other problems (Moore et al., 2018), to the extent that children diagnosed with APD do not, on average, perform differently on many listening, language and cognitive tests from children assessed to have other neurodevelopmental conditions^1^ (Dawes & Bishop, 2009; de Wit et al., 2018; Ferguson, Hall, Riley, & Moore, 2011; Sharma, Purdy, & Kelly, 2009). A long-standing issue is that, due to lack of a solid theoretical basis, established physiological mechanisms, or consensus on the definition of APD, appropriate diagnostic testing, and intervention cannot be rationalized (Papesh, Fowler, Pesa, & Frederick, 2023; Vermiglio, 2014). LiD, by contrast, focuses on symptoms commonly reported by the caregivers of children who present at audiology clinics, but have clinically normal audiograms (Barry & Moore, 2021; Dillon & Cameron, 2021; Petley et al., 2021; Shiels, Tomlin, & Rance, 2023). LiD allows the underlying causes to be multifactorial and not necessarily due to auditory system dysfunction. APD is not a “clinical entity”, as defined by the Sydenham-Guttentag criteria (Vermiglio, 2014), whereas self- or caregiver report, the basis of LiD, may be a more ecologically valid reference standard for diagnostic category (Vermiglio, Soli, & Fang, 2018). Having stated these definitions and distinctions, the term LiD is preferentially used throughout this paper except where APD is specifically referred to in a previous diagnosis or a reference.

Interventions for LiD in children include hearing aids, wireless, remote microphone hearing devices (Johnston, John, Kreisman, Hall, & Crandell, 2009; Lemos, 2009; Shiels et al., 2023), aural rehabilitation (Boothroyd, 2007), usually in the form of computer game-based training (e.g. Fast ForWord; (Strong, Torgerson, Torgerson, & Hulme, 2011), “metacognitive training” (Chermak & Musiek, 2013), classroom accommodations (DeBonis, 2015), and other listening exercises including dichotic listening, language and sound discrimination training (Fey et al., 2011; Lotfi Y, 2016; Moncrieff, Keith, Abramson, & Swann, 2017; Sharma, Purdy, & Kelly, 2012). The level of evidence for the effectiveness of these interventions is variable but, in general, does not support their widespread clinical adoption (Fey et al., 2011). One series of studies found, however, that training using a spatialized sound task (LiSN & Learn^2^) resulted in improved spatial hearing for children identified with ‘spatial processing disorder’ (Cameron & Dillon, 2011). A gold-standard, blinded control method, and independently developed self- and caregiver-report questionnaires, suggested in subsequent studies that more generalized aspects of listening may also be improved (Cameron, Glyde, & Dillon, 2012; Cameron, Glyde, Dillon, King, & Gillies, 2015). Recently, (Shiels et al., 2023) provided strong evidence for the effectiveness of remote microphone devices in aiding LiD. To date, however, no intervention method has seen general clinical acceptance. High quality, large-scale clinical trials have not been performed and there is no consensus on what outcome measures to use. Consequently, LiD is a challenging condition for parents, providers, and insurers to understand, evaluate, endorse, and fund.

To address questions about the underlying mechanisms of childhood LiD, Cincinnati Children’s Research Foundation (CCRF) and the National Institute of Deafness and Communication Disorders (NIDCD), funded a longitudinal project, Sensitive Indicators of Childhood Listening Difficulties (SICLiD). Children (6-13 years old) with clinically normal audiometry were recruited into two groups, listening difficulty (LiD) or typically developing (TD), based primarily on the results of a validated and reliable caregiver evaluation instrument (the ECLiPS; Barry & Moore, 2021; Denys et al., 2024; Petley et al., 2021). A key premise of the ECLiPS is that caregivers, typically parents, observe their children’s behavior over extended periods in everyday life and are therefore best able to judge the child’s listening and related abilities. A similar rationale was used to develop a widely-used caregiver report of children’s language abilities (CCC-2; Bishop, 2006) that inspired the ECLiPS design. The ECLiPS has 38 statements (items) that various stakeholder groups suggested are characteristics of children with LiD and that the caregiver rates on a 5-point Likert scale from ‘strongly agree’ to ‘strongly disagree’. Children assigned to both TD and LiD groups were tested on a range of audiological and other physiological and behavioral measures. Papers published to date have focused on the results of those tests (Hunter et al., 2023; Hunter et al., 2021; Moore, Hugdahl, Stewart, Vannest, Perdew, Sloat, Cash, & Hunter, 2020; Petley et al., 2024; Petley et al., 2021; Stewart et al., 2022). They suggest that LiD/APD is a result primarily of widespread impaired cognitive processes (e.g. attention, memory, executive function), and speech-language processing mechanisms beyond the central auditory nervous system.

The families of children with LiD who participated in the SICLiD project were approached to consent for their child’s relevant medical history to be accessed by this study’s investigators. The overall goal of that access, and of the study reported here, was to investigate the hearing and related assessments and interventions the children received within the clinical divisions of Cincinnati Children’s Hospital Medical Center (CCHMC) in relation to the research measures in the same children. Nearly all the clinical assessments and interventions had been provided before the children entered the SICLiD study and, in all cases, the research results were unknown to the clinical providers. We hypothesized, based on previous studies (Moore et al., 2018), that the children would have received a wide variety of clinical assessments, but relatively few and diverse interventions (Fey et al., 2011). Specific questions asked here were: What provider type and how many clinical assessments were given? How did the records of those clinical assessments relate to the research results of the SICLiD study? What type of interventions have been used?

## Methods

### Participants and caregiver reports

Participants were all 74 children, aged 6-13 years, who completed the SICLiD research study baseline assessment and had significant LiD as defined by Total ECLiPS score more than one s.d. below^3^ the mean TD score (Petley et al., 2021). Caregivers of all recruited children also completed the CCC-2 (Bishop, 2006), and a background questionnaire concerning demographics (age, race/ethnicity, caregiver education level), history of neurologic and otologic appointments, and diagnoses or assessments of learning disorders. Those with severe neurologic, otologic, or intellectual conditions (e.g. current use of tympanic pressure equalizing, PE, tubes, unable to complete tasks) were excluded, but those with common neurodevelopmental disorders known to overlap with LiD (e.g. language, attention, executive function, high-functioning autism spectrum disorder - ASD, history of otitis media) were included.

### Hearing and cognitive tests

Both clinical testing and research were conducted within CCHMC. All 74 children had an extensive audiometric assessment (Hunter et al., 2021), including pure tone audiograms (octave frequencies from 0.25 – 8.0 kHz, plus 10, 12.5, and 16 kHz, bilaterally), wideband tympanometry, acoustic reflex growth, transient and distortion product otoacoustic emissions, as well as auditory brainstem and cortical evoked responses (Hunter et al., 2023; Petley et al., 2024). In other behavioral assessments, standardized scores on the SCAN-3:C (Keith, 2009), the test suite most commonly used in the US to diagnose APD (Emanuel, Ficca, & Korczak, 2011), and on all 4 SCAN subtests, were obtained. Standardized scores on the Low- and High-Cue, and the Spatial, Talker and Total Advantage subtests of the Listening in Spatialized Noise: Sentences Test (LiSN-S; Cameron & Dillon, 2007), and on all subtests and Composites of the National Institutes of Health (NIH) Cognition Toolbox (Weintraub et al., 2013) were also obtained (see Petley et al., 2021, for further details). Performance on the Bergen Dichotic Listening Test (DL; Hugdahl et al., 2009; Moore et al., 2020) was available for a sub-set (n=44) of the children.

### Hospital records

For the 64/74 children with LiD who had an electronic medical record (Epic) that included clinical assessments of hearing, listening, and/or other developmental disorders prior to the SICLiD study, a retrospective analysis of each child’s record was completed. Each service operated differently with respect to the clinical assessment. The assessment result was often not clearly documented, and many different terms were used to describe the child’s identified problems and intervention recommendations. For this reason, we are using the term “condition” rather than “diagnosis”, to categorize the problems identified for each participant. One quantifiable difference was the age at first assessment, that varied from 1-12 y.o., with speech delays and otitis media typically recorded early and behavioral disorders recorded at school age. For most cases, there was a history of assessments spanning several years, often right up to the time of first participation in the SICLiD study.

For each participant, a summary description of conditions related to APD or LiD was first noted. Then, for each clinical Division - Speech-Language Pathology (SLP), Audiology, Developmental and Behavioral Pediatrics (Psychology^4^), Physical and Occupational Therapy (OT), and for each participant, we extracted and coded the following data: appointment type (assessment, therapy), goals, number of encounters, progress/outcome. Assessment and therapy types were coded (up to 6 types per Division for each child). Multiple authors independently performed repeat summaries and counts of the EPIC records. These included research coordinators (RB, JC-K) and all work was overseen by senior authors (LLH, DRM) who also made complete, independent assessments of the records and the final decision on how to categorize each child’s records. All authors coded the clinical records without knowledge of each child’s performance in the SICLiD study.

#### Analysis

Caregiver, hearing, listening, and cognitive scores were presented as charts or violin plots. Violin plots showed probability density of the data, and median, interquartile range and total range in the context of standardized scores expected from TD children.

Epic clinical assessment data were manually coded and reduced to five summary conditions: Language (including speech, vocabulary, reading, apraxia/dyspraxia, stuttering), Hearing (including APD/LiD, sensory processing^5^, hearing loss^6^), Attention (including attention deficit hyperactivity disorder, ADHD), Anxiety (including panic, stress, post-traumatic stress disorder, PTSD, and depression), and ASD (including social or pragmatic problems). Each child was assigned a binary code (0 – no symptoms, 1 – symptoms; Table 1) for each condition.

**Table 1.**
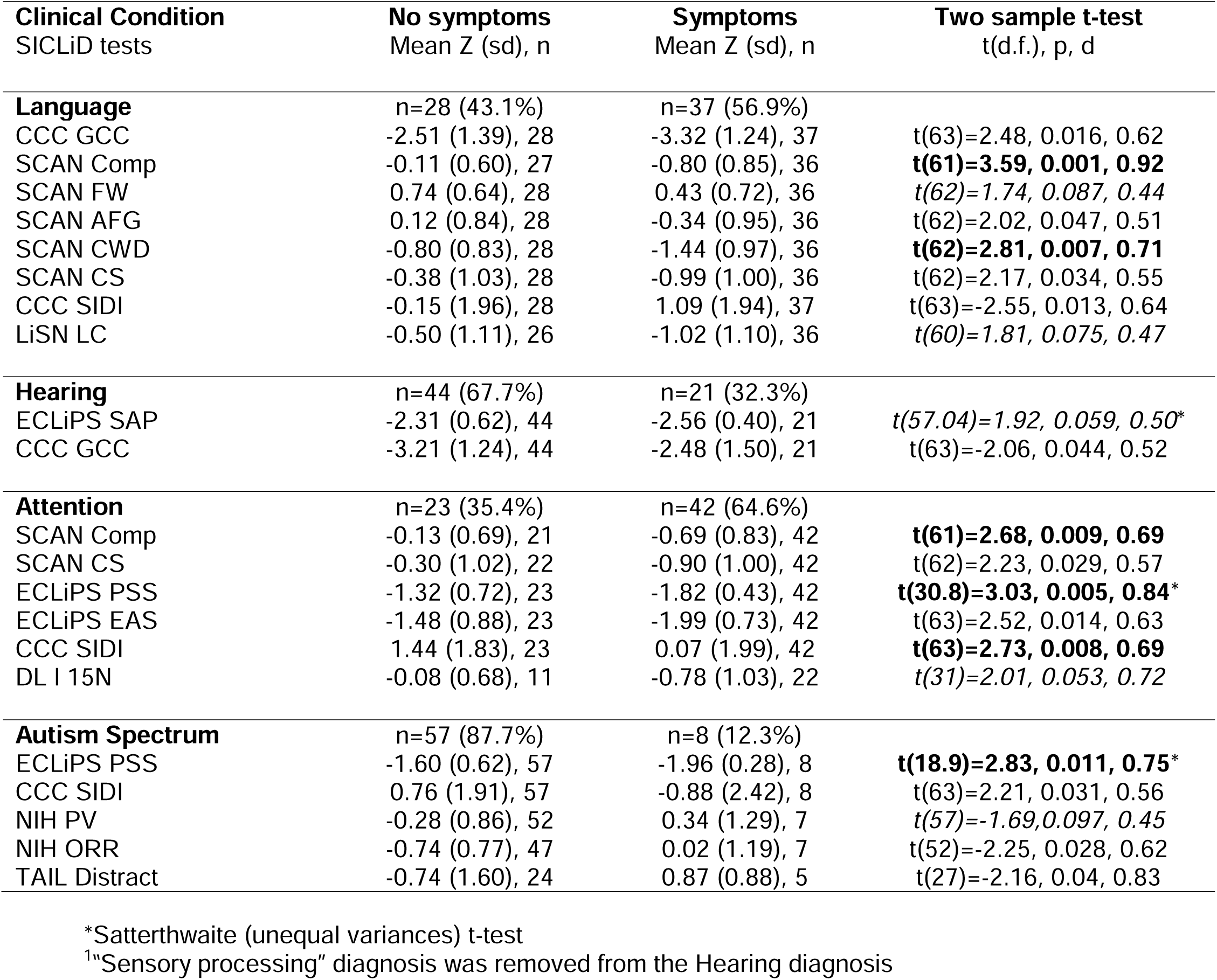
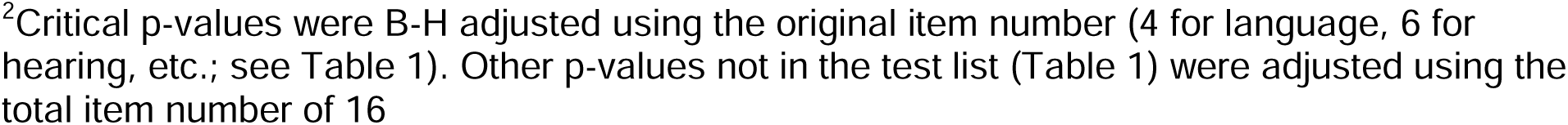
Significant SICLiD predictor tests of clinical assessments. Univariate analysis. Assessment clinical conditions (labels in bold) were Language, Hearing (and Processing), Attention, ASD and APD, although no significant predictors for APD were found. For each condition, the number of children with LiD, but without or with symptoms of that condition, is shown in the top row. Following rows show SICLID test results (e.g. CCC GCC; see Abbreviations), scaled z-score means, number of children (n), and two sample t-test results (far right column) with probability (p) and effect size (d). (*italics*: 0.05 < p < 0.1, **bold**: p < 0.01.).

| **Clinical Condition**<br>SICLiD tests | **No symptoms**<br>Mean Z (sd), n | **Symptoms**<br>Mean Z (sd), n | **Two sample t-test**<br>t(d.f.), p, d |
| --- | --- | --- | --- |
| **Language** | n=28 (43.1%) | n=37 (56.9%) |  |
| CCC GCC | -2.51 (1.39), 28 | -3.32 (1.24), 37 | t(63)=2.48, 0.016, 0.62 |
| SCAN Comp | -0.11 (0.60), 27 | -0.80 (0.85), 36 | **t(61)=3.59, 0.001, 0.92** |
| SCAN FW | 0.74 (0.64), 28 | 0.43 (0.72), 36 | t(62)=1.74, 0.087, 0.44 |
| SCAN AFG | 0.12 (0.84), 28 | -0.34 (0.95), 36 | t(62)=2.02, 0.047, 0.51 |
| SCAN CWD | -0.80 (0.83), 28 | -1.44 (0.97), 36 | **t(62)=2.81, 0.007, 0.71** |
| SCAN CS | -0.38 (1.03), 28 | -0.99 (1.00), 36 | t(62)=2.17, 0.034, 0.55 |
| CCC SIDI | -0.15 (1.96), 28 | 1.09 (1.94), 37 | t(63)=-2.55, 0.013, 0.64 |
| LiSN LC | -0.50 (1.11), 26 | -1.02 (1.10), 36 | t(60)=1.81, 0.075, 0.47 |
| **Hearing** | n=44 (67.7%) | n=21 (32.3%) |  |
| ECLiPS SAP | -2.31 (0.62), 44 | -2.56 (0.40), 21 | t(57.04)=1.92, 0.059, 0.50* |
| CCC GCC | -3.21 (1.24), 44 | -2.48 (1.50), 21 | t(63)=-2.06, 0.044, 0.52 |
| **Attention** | n=23 (35.4%) | n=42 (64.6%) |  |
| SCAN Comp | -0.13 (0.69), 21 | -0.69 (0.83), 42 | **t(61)=2.68, 0.009, 0.69** |
| SCAN CS | -0.30 (1.02), 22 | -0.90 (1.00), 42 | t(62)=2.23, 0.029, 0.57 |
| ECLiPS PSS | -1.32 (0.72), 23 | -1.82 (0.43), 42 | **t(30.8)=3.03, 0.005, 0.84\*** |
| ECLiPS EAS | -1.48 (0.88), 23 | -1.99 (0.73), 42 | t(63)=2.52, 0.014, 0.63 |
| CCC SIDI | 1.44 (1.83), 23 | 0.07 (1.99), 42 | **t(63)=2.73, 0.008, 0.69** |
| DL I 15N | -0.08 (0.68), 11 | -0.78 (1.03), 22 | t(31)=2.01, 0.053, 0.72 |
| **Autism Spectrum** | n=57 (87.7%) | n=8 (12.3%) |  |
| ECLiPS PSS | -1.60 (0.62), 57 | -1.96 (0.28), 8 | **t(18.9)=2.83, 0.011, 0.75\*** |
| CCC SIDI | 0.76 (1.91), 57 | -0.88 (2.42), 8 | t(63)=2.21, 0.031, 0.56 |
| NIH PV | -0.28 (0.86), 52 | 0.34 (1.29), 7 | t(57)=-1.69, 0.097, 0.45 |
| NIH ORR | -0.74 (0.77), 47 | 0.02 (1.19), 7 | t(52)=-2.25, 0.028, 0.62 |
| TAIL Distract | -0.74 (1.60), 24 | 0.87 (0.88), 5 | t(27)=-2.16, 0.04, 0.83 |

\*Satterthwaite (unequal variances) t-test
<sup>1</sup>“Sensory processing” diagnosis was removed from the Hearing diagnosis
<sup>2</sup>Critical p-values were B-H adjusted using the original item number (4 for language, 6 for hearing, etc.; see Table 1). Other p-values not in the test list (Table 1) were adjusted using the total item number of 16

All SICLiD scores were converted to age-specific Z-scores so that the relation between each score was equalized to ascertain prediction level for each clinical condition. Mean Z-scores were first compared between children without or with symptoms of each condition (0 or 1 above) using two sample or Satterthwaite t-tests to assess the univariate statistical significance (p-value) and effect size (d) of the difference (Table 1). Critical p-values were assessed using Benjamini-Hochberg adjustments to determine a false discovery rate < 5%. However, because of the relatively small number of SICLiD tests that were predictors, additional tests with marginal statistical significance (0.05 < p < 0.1) were also included in Table 1. Data of children assessed for APD by the CCH Audiology Department prior to the SICLiD study were, additionally, analyzed separately using multiple t tests comparing scores on all SICLiD measures between children with (n = 22) and without (n = 22) an APD diagnosis (including auditory processing ‘weakness’; see (D.R. Moore et al., 2018). ECLiPS Total score was the independent variable throughout.

SICLiD univariate predictors listed in Table 1 were considered in a subsequent adjusted logistic regression model of clinical assessments. Relationships and multicollinearity among the predictors were explored using the correlation coefficient and variance inflation factor (VIF). A high correlation and a VIF greater than 5 indicates problematic multicollinearity. The final logistic prediction models (Table 2) were selected with the best prediction combinations of SICLiD scores, reported with odds ratios (with 95% confidence intervals) and an area under the receiver operating characteristic (AUROC) curve. A recent review of the clinical AUROC literature warned against labeling results (e.g. ‘good discrimination’; (de Hond, Steyerberg, & van Calster, 2022), so only AUROC values are presented here. Data analysis was conducted using SAS statistical software, version 9.4 (SAS Institute, Cary, N.C.). The research described here was approved by the Cincinnati Children’s Hospital Research Foundation Institutional Review Board.

**Table 2.**
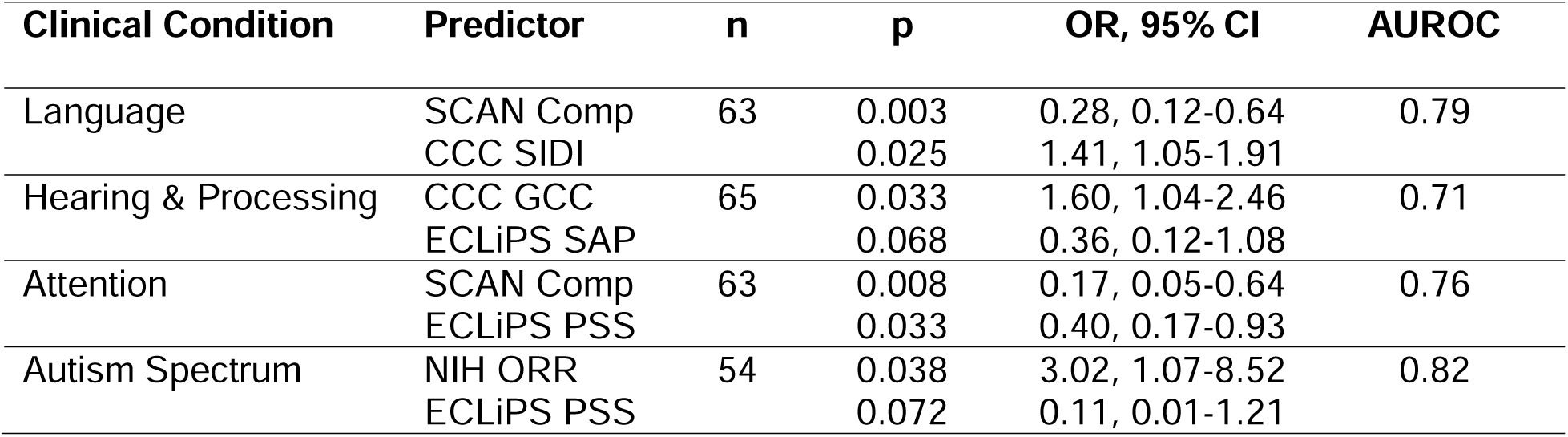
Best logistic models for SICLiD research predictors of clinical conditions. The first named predictor for each condition had the greater discriminatory power, indexed by the predictor probability (p). ‘n’ is the number of participants completing both predictor tasks, ‘OR’ is the odds ratio (with 95% confidence intervals) for each predictor, AUROC values show the final discriminatory power for each model (scale 0.5 – 1.0). Further details in the Table 1 caption and in the text.

| **Clinical Condition** | **Predictor** | **n** | **p** | **OR, 95% CI** | **AUROC** |
| --- | --- | --- | --- | --- | --- |
| Language | SCAN Comp | 63 | 0.003 | 0.28, 0.12-0.64 | 0.79 |
|  | CCC SIDI |  | 0.025 | 1.41, 1.05-1.91 |  |
| Hearing & Processing | CCC GCC | 65 | 0.033 | 1.60, 1.04-2.46 | 0.71 |
|  | ECLiPS SAP |  | 0.068 | 0.36, 0.12-1.08 |  |
| Attention | SCAN Comp | 63 | 0.008 | 0.17, 0.05-0.64 | 0.76 |
|  | ECLiPS PSS |  | 0.033 | 0.40, 0.17-0.93 |  |
| Autism Spectrum | NIH ORR | 54 | 0.038 | 3.02, 1.07-8.52 | 0.82 |
|  | ECLiPS PSS |  | 0.072 | 0.11, 0.01-1.21 |  |

## Results

### Research tests of hearing, listening and cognition

Performance on SICLiD tests of hearing, listening, and cognitive function for this sample of children with LiD is summarized in Fig. 1. Performance of the entire SICLiD sample (n = 146) on behavioral tests is reported in detail elsewhere (Hunter et al., 2021; Kojima, 2024; Petley et al., 2021). All participants with LiD had clinically normal hearing, including pure tone audiograms (PTA ≤ 20 dB HL bilaterally at each octave frequency from 0.25 – 8.0 kHz). Additional, extended high frequency testing (10, 12.5, and 16 kHz, bilaterally) revealed substantial variability, with some hyper-acute (−15 dB HL) and insensitive (to 50 dB HL) thresholds (Fig. 1A). Total ECLiPS scores were uniformly low, since this was the primary inclusion criterion. ECLiPS scales (SAP etc.) were more variable than the Total, but all had a median score between 3-5, well below the median standard score of 10 (Fig. 1B, D). CCC-2 “general communication composite” (GCC) scores correlated with Total ECLiPS scores (r = 0.39; p < 0.001) and revealed associated language problems of these children (Figs. 1B,C). CCC-SIDI scores (Fig. 1C) were above the typical range (> +10) for 23/74 participants, suggestive of a pragmatic language impairment, and below typical (< +10) for 9/74 participants, suggestive of ASD (Bishop, 2003). The SCAN suite of tests showed generally poor performance on two tests of dichotic listening, the Competing Words and Competing Sentences, average performance (10) on a speech-in-noise test (Auditory Figure-Ground) and above average performance on a test of low-pass Filtered Words (Fig. 1E). The Listening in Spatialized Noise – Sentences (LiSN-S) showed that children with LiD generally performed poorly in this complex, competing speech test. This was the case for both individual tests (low cue, high cue), and for the Spatial Advantage and Talker Advantage derived measures, where cognitive aspects of performance are minimized (Fig. 1F; see (Petley et al., 2021), but not for the Total Advantage derived measure. A suite of cognitive tests, the NIH Cognition Toolbox (Weintraub et al., 2013), showed widespread and relatively uniform difficulty for performance on an array of mostly visual-based cognitive tasks (Fig. 1G). Performance on the various conditions of the Bergen DL (Moore et al., 2020) did not show any difference between groups, with one exception, in the condition (ILD = −15 dB) for which laterality was more pronounced in the LiD group. Performance differences between children with and without diagnosed APD on all SICLiD tests (n = 28) were non-significant (p > 0.05).

**Figure 1:**
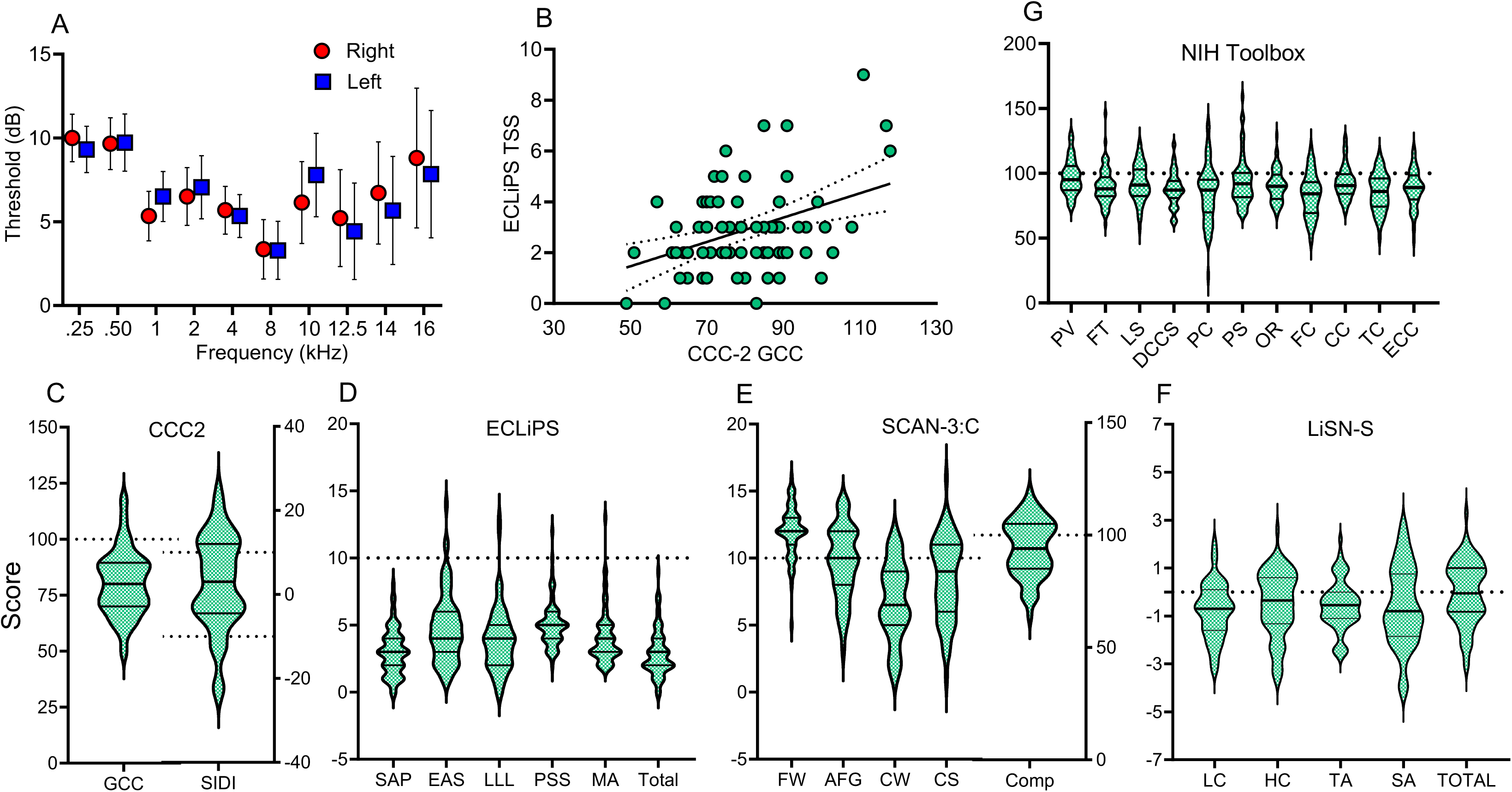
Auditory and cognitive SICLiD test scores of children enrolled in the study. A. Mean (± 95% CI) audiometric thresholds. B. Individual ECLiPS Total scaled score (mean typical = 10) and CCC GCC (mean typical = 100) caregiver report scaled scores with linear best fit. C. Scaled score density (violin plots in panels C-G) for the CCC GCC (left axis) and CCC SIDI (right axis; typical range −10 to 10). D. ECLiPS subscale (SAP etc) and Total scaled scores (mean typical = 10). E. SCAN-3:C subscale (FW etc, mean typical = 10) and Composite (mean typical = 100) scaled scores. F. LiSN-S subscale (LC etc) and Total Advantage scaled z-scores (mean typical = 0). G. NIH Cognition Toolbox tests (PV – OR), Fluid composite (FC), Crystalized composite (CC), Total composite (TC), Early Childhood composite (ECC). Mean typical = 100. Further details in Methods.

### Clinical encounters and assessments

Number of encounters and assessments for each Hospital service is shown in Fig. 2. Ten of the children did not have a clinical record at CCHMC, although the caregiver background questionnaires of at least three suggested they had been assessed elsewhere with an attention or speech/language problem. Of the remaining 64, the mean number of services used by each participant was 2.0. 75% attended at least one appointment with Audiology, and (69%) with SLP, while 28% were seen by Psychology and 30% by OT (Fig. 2A). A total of 130 assessments of problem conditions of Attention, Language, Hearing (& ‘Processing’), Anxiety, and ASD were made among the sample of 64 children (see Supplementary Results for details), a mean of 2.03 conditions per child. The most common condition was Attention, followed by Language, Hearing, Anxiety and high-functioning ASD. Two children had been assessed with all 5 conditions coded here (Fig. 2E). Aside from an overall Hearing condition, of whom 88% had an APD diagnosis, the balance of Language, Attention, and ASD between the two groups with or without an APD diagnosis was near identical. However, more than twice as many children without APD had an Anxiety assessment as those with APD (Figs. 2G,H). This difference was reduced when the three children with sensory processing disorder were removed (Fig. 2F). Anxiety is not considered further here as we did not have any SICLiD tests for it.

**Figure 2:**
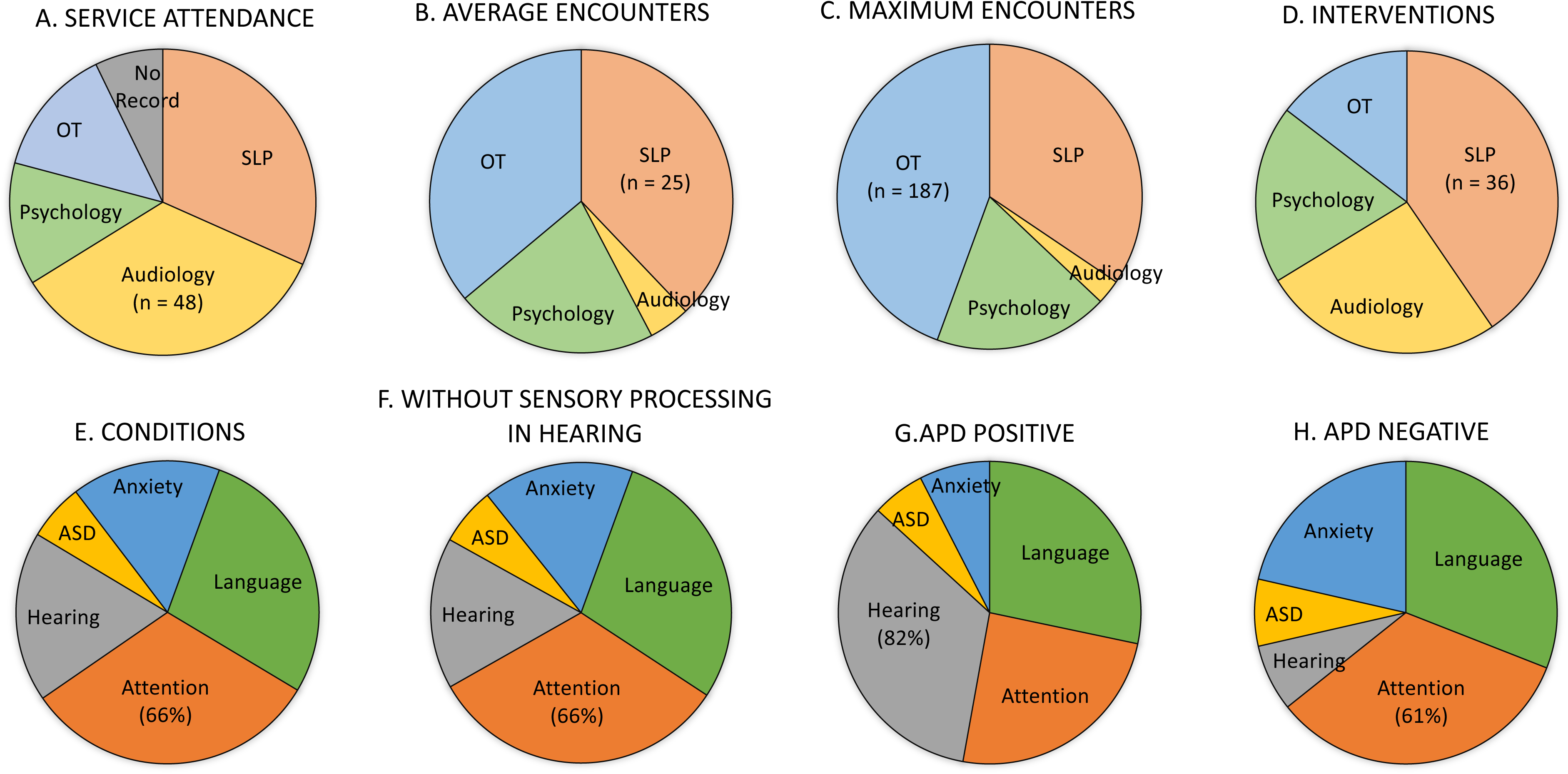
Children attending each clinical service (A – D) and assessed for each disorder (E – H). For each pie chart, the largest slice is labeled with the number of participants (n) or the percentage of all participants (%). Note that percentages did not add to 100 because many children had two or more assessments.

### Clinical Intervention and outcomes

Although there is no consensus in the literature on what treatment children with LiD should be offered, we document here the follow-up that occurred in this sample. Interventions were highly heterogenous, and not consistently documented in the clinical records. The number of different interventions in SLP clinics (n = 36) was about twice that in any of the other three clinics (Fig. 2D; Supplementary Results Tables S3 – S6). Some services recommended interventions to most of the children they saw (Psychology – 94%, SLP – 82%, OT – 68%), while Audiology recommended interventions to fewer than half the children they saw (48%). Audiology recommended interventions were most commonly behavioral (in order: accommodations, computer training, communication strategies; Supplementary Results Table S7), but devices, specifically remote microphone and/or low-gain hearing aids, were also popular. For the other services, a broad range of exclusively behavioral interventions was used. Most strikingly, both the average (Fig. 2B) and maximum (Fig. 2C) number of encounter sessions varied hugely between the specialties, with SLP (mean = 25) and OT (max = 187) having many intervention sessions, Psychology near the middle, and Audiology (mean = 3) having a small number of encounters that were mostly assessment. It was difficult to separate the intervention from the assessment, as these were often not specified separately. This challenge was particularly apparent in SLP, where the assessment and intervention often seemed to be one and the same thing. For example, assessment: “speech disturbance, articulation (/s/,/f/,/sh/,/z/), voice disturbance”, intervention: “work on speech, articulation and voice disturbances.”

It was often unclear whether the children participated in the stated interventions, or whether the interventions were only suggestions of procedures that could be tried. All the services except Audiology provided behavioral interventions during the encounter, whereas Audiology tended not to intervene for children with LiD but with clinically normal audiograms.

Scant evidence was available on outcomes, with almost no evidence of follow-up. However, SLP had the most consistent documentation on cessation of therapy. That service also had by far the largest number of different specified interventions (n = 35 among 44 children), and the highest mean number of intervention sessions (24.9).

### Relation between clinical assessments and research test scores

Following the methods described above, the prediction value for each clinical condition by each SICLiD research test was assessed. Of 80 univariate comparisons, only 16 showed significant differences (adjusted 17 < 0.05) between research scores based on binary clinical assessment category (i.e. without or with symptoms) and an effect size ≥ 0.5. An additional 5 comparisons with marginal statistical significance (0.1 > p > 0.05), two of which had p ≥ 0.5, are included in Table 1. Of these 21 comparisons, 9 were from the caregiver report measures (ECLiPS, CCC-2), 7 were from the SCAN, and 3 were from the NIH Oral reading test. Among conditions, Language, Attention, and Autism Spectrum had the largest numbers of predictive test scores, whereas the SCAN Composite score had the highest level of predictability (d = 0.92; for Language) of any single test.

Overall, clinical conditions were largely independent of research test scores, with the most predictive research scores for clinical groups coming from the language-centered caregiver report, the CCC-2. The CCC-2 also predicted clinical Language problems well, but not as well as the SCAN Composite, designed as a test for APD. For clinical Hearing and Processing, on the other hand, the caregiver ECLiPS SAP and CCC GCC were both moderately predictive, as expected. But none of the SCAN tests, nor any of the other SICLiD measures of speech-in-noise, cognition, or dichotic listening were significantly predictive of Hearing and Processing.

The caregiver ECLiPS PSS and CCC SIDI, both reporting aspects of pragmatic language, also predicted clinical Attention and ASD-like symptoms, with the SCAN Composite and Competing Sentences (SCAN CS subtest) also predicting Attention. The NIH Oral Reading Recognition (ORR) and Picture Vocabulary (PV) Tests, designed primarily for measuring aspects of Language, and the TAIL Distraction Test, designed for measuring Attention, were all predictive of ASD symptoms. Children previously assessed for APD and either diagnosed (n=22) or not diagnosed (n=22) did not show any significant or near-significant differences on performance of any of the research tasks related to their diagnostic status. Consequently, no significant APD research predictors were identified.

### Predictive assessment logistic models

Combining two or more research tests might provide better prediction of a clinical condition than just a single test. In this final analysis we created logistic regression models for each clinical condition in which at least marginal significant research predictors had been identified in the univariate analysis (Table 1). In all cases, the final models provided higher predictability than individual tests.

#### Language

The final prediction model consisted of the SCAN Composite and CCC SIDI (Table 2). SCAN Composite subscales (FW, AFG, CWD, CS) had high multicollinearity, with VIFs > 30 (see Methods). CCC GCC and LiSN LC subscale were highly correlated with the SCAN Composite (both p < 0.001). SCAN and LiSN subscales, and CCC GCC were therefore removed from the final model.

#### Hearing and Processing

The final model included the same predictors identified in the univariate analysis - CCC GCC and ECLiPS SAP (Table 2). Since the correlation between these scaled predictors of Hearing and Processing was non-significant, collinearity was not an issue.

#### Attention

The SCAN Comp was, again, a strong predictor of attention and achieved a relatively high AUROC when paired with the ECLiPS PSS (Table 2). Other potential pairings between these predictors and the ECLiPS EAS, the DLI15N and the CCC SIDI were rejected because of lower effect size, small sample size, and wide confidence intervals. The ECLiPS EAS also correlated highly with the ECLiPS PSS, although none of the predictors had VIF > 3.

#### Autism Spectrum Disorder

The final prediction model included NIH OR and ECLiPS PSS, yielding a high AUROC (Table 2). No correlations between ASD predictors exceeded 0.5 and all had VIF < 1.5. Other combinations, including the TAIL Distract, also yielded impressive ASD prediction, but the TAIL sample size was small.

## Discussion

In this study we found that quantitative research measures of LiD and APD were generally not good predictors of clinical assessments or interventions when used in isolation. Caregiver report scores were more aligned with clinical conditions and, when used in conjunction with objective measures, provided reasonable prediction.

Nearly all children with LiD recruited into the SICLiD study had previous encounters with clinical specialists working in audiology and related professions; 86% had attended relevant services at CCHMC, and caregiver reports for the remainder reported other professional interactions. As found in other, independent samples, and hypothesized here, most children had been seen by more than one type of clinical specialist due to multiple behavioral problems observed by caregivers (Ferguson et al., 2011; Moore et al., 2018; Sharma et al., 2009). Families of children with LiD thus have a wide range of concerns that extend well beyond the audiological domain. Our other hypothesis, that few, diverse interventions would be used, as previously found for audiology (Emanuel et al., 2011), was partly supported. For Audiology, the previous finding was confirmed, with fewer children than in other services receiving any recommended intervention. For the other services, most children received behavioral intervention recommendations that were delivered in serial therapy sessions. Interventions may have been low for Audiology because that profession is device oriented, rather than therapy focused. Perhaps related to this, many audiologists believe that they should diagnose APD, and that SLPs should provide behavioral therapy for it (Emanuel et al., 2011). This may be appropriate, since increasing evidence suggests that APD may be specific to various aspects of speech and language synthesis and understanding (Petley et al., 2021; Stewart et al., 2022), such as reading (Magimairaj, Nagaraj, Sergeev, & Benafield, 2020).

### Relations between lab and clinical data

We found that clinical assessments, and the intervention suggestions that followed, showed little overall relationship to the research results of the SICLiD study. The most striking difference between children with and without LiD in the SICLiD study was the consistently poorer performance of the children with LiD across the NIH Cognition Toolbox tests (Petley et al., 2021), supporting the hypothesis that LiD is primarily a cognitive impairment, rather than an auditory sensory problem (de Wit et al., 2018; Moore, Ferguson, Edmondson-Jones, Ratib, & Riley, 2010; Petley et al., 2021; Tomlin, Dillon, Sharma, & Rance, 2015). In this study, however, performance on only two language tests (NIH ORR, NIH PV), among the seven NIH Toolbox subtests examined, predicted children with just a single clinical condition (ASD). We cannot explain this discrepancy. One possibility lies in our recent finding, from the longitudinal SICLiD data, that maternal education level and spatial listening skills, as well as cognitive skills, are independently predictive of the degree of LiD (Kojima et al., 2024). It may be that, in the context of clinical assessment, these other factors were more influential than they were in the research setting. This seems particularly plausible for maternal education, a proxy for socioeconomic status. For this or other reasons, clinicians may look beyond a child’s overall cognitive capacity during more target-directed neurodevelopmental assessment.

Speech-in-noise listening has been suggested as a common problem, or even *the* problem experienced by children with LiD (Audiology, 2010; Cameron et al., 2015; Campbell et al., 2019; Petley et al., 2021; Saunders & Haggard, 1989; Sharma, Dhamani, Leung, & Carlile, 2014; Vermiglio, 2014). It was therefore surprising that, in this study, we did not find a single significant relationship between three subtests of the LiSN-S and the five clinical conditions. A near-miss was the Low Cue subtest (LiSN LC) that predicted (p=0.08; d=0.47) children with a language problem, but no research tests were predictive of APD. The general lack of relationship with the clinical conditions suggests that the LiSN-S, a test we chose primarily because of its likely relevance to everyday communication in noisy environments, may not capture well the everyday problems that children with LiD experience or, at least, relate to clinical observations. Based on these findings, we predict that other commonly used speech-in-noise tests (see Billings, Olsen, Charney, Madsen, & Holmes, 2024, for a review) may also be unable to capture those problems.

### Caregiver reports

Caregiver reports (ECLiPS, CCC-2) accounted for half of the significant predictions by SICLiD tests of clinical assessments in this study. Caregiver reports were thus better predictors of overall clinical assessments than the research tests of hearing and cognition used in SICLiD. Previously, (Bishop, Laws, Adams, & Norbury, 2006) and (Bishop & McDonald, 2009) showed that CCC-2 was as good a predictor of clinically assessed language impairment as widely used, standardized tests, such as the NEPSY suite for speech repetition ability (Korkman, 1998) and other tests of reading ability (Neale, 1997; Torgeson, Wagner, & Rashotte, 1999). Here, we also found that the CCC-2 identified clinical language problems well. Also surprising was that children with a clinically identified Hearing & Processing condition, most of whom were diagnosed with APD, were predicted *only* by the caregiver report measures. None of the other SICLiD measures, such as speech-in-noise (LiSN-S), cognitive (NIH Toolbox), or dichotic listening ability (SCAN CW, SCAN CS, DL), frequently associated with APD in the literature (Bamiou, Musiek, & Luxon, 2001; de Wit et al., 2018; Dillon & Cameron, 2021; Moore et al., 2010; Sidiras, Iliadou, Chermak, & Nimatoudis, 2016), predicted clinical Hearing & Processing conditions.

These apparent mismatches between research and clinical findings, and the diagnostic associations with which they are assumed to be directed, were not limited to Language and Hearing & Processing, however. Both caregiver reports and the SCAN Comp also predicted clinical Attention, while standardized tests of reading and attention, together with the ECLiPS caregiver report PSS subscale, most strongly predicted ASD. Concerning Attention, the SCAN Comp (speech-based auditory), ECLiPS PSS (caregiver rated pragmatic and social skills) and the CCC SIDI (caregiver rated social interaction) had the largest effect sizes. The SCAN is discussed further below. The significant role of attention as a predictor of social awareness reflects the real-world observations of caregivers and clinicians and may an important area for further research. The general lack of discriminative relationship between clinical assessments and current, standard test measures may reflect other different perceptions between neurodevelopmental scientists and clinicians. In response, adult hearing research is pivoting towards more ecological evaluations of everyday listening environments, using techniques such as ecological momentary assessment (Holube, von Gablenz, & Bitzer, 2020) and large-scale, electronic health record assessments of diverse populations (Saunders, Dillard, Zobay, Cannon, & Naylor, 2021). Adaptations of these techniques may also provide valuable data for pediatric populations.

### Predicting clinical assessments

Bishop & McDonald (2009) found that combining the results of a language test (NEPSY) with those of the CCC-2 led to the best discrimination between clinically referred and non-referred children, suggesting that these assessment approaches provided useful complementary information. We therefore asked whether multiple test scores might yield better prediction of the clinical conditions more generally and, indeed, that was the case. Across assessments, pairing tests represented the best compromise between minimizing collinearity, larger sample and effect sizes, and maximizing AUROC values. The end results were impressive, suggesting that the SCAN Composite, along with caregiver report measures (CCC and ECLiPS) and, for ASD, the NIH Toolbox Oral Reading Recognition test were more predictive of clinical assessments than any individual measures. Again, the SCAN, especially the dichotic tests (CWD, CS), was very sensitive to Attention and Language, particularly when used together with caregiver reports.

The SCAN battery is the most used clinical test for diagnosing APD in the U.S. (Emanuel et al., 2011). However, in this study it was a better predictor of clinical attention and language problems than of auditory problems. One possible explanation for this apparent contradiction is that it was designed in the clinic, by clinicians, with inattentive and language-challenged children as the main target groups. The SCAN is made up of suprathreshold, fixed-level, speech-based tasks rather than adaptive SiN tests that are better predictors of peripheral hearing sensitivity (such as digits-in-noise; Smits et al., 2013). Thus, the SCAN may operate most sensitively at higher levels of processing that tap attention and language skills. The CWD and CS SCAN subtests, like SiN tests, also require repetition of words and sentences (language skills), but in a dichotic format, where different words or sentences are delivered to each ear. Such tests led to the influential Filter Theory of selective attention (Broadbent, 1958). Subsequent research showed that the filter was “leaky” (Egeth, 1992), for example letting speech with salient meaningfulness, such as a person’s own name, through the filter (Cherry, 1953). Although acoustic dissimilarities between dichotic stimuli contribute to response recall accuracy (Musiek & Chermak, 2015), the finding that semantic cues contribute to performance in individuals with normal audiograms suggests a level of processing beyond the classical central auditory system.

### Limitations and further work

Responses to caregiver report items in the study reported here may have reflected the (earlier) clinical assessments and may thus not have been a fair measure of association. However, the time lag between clinical visits and research testing was typically months to years, and the items in the ECLiPS and CCC-2 are, for the most part, not obviously related to specific clinical assessments.

Two SICLiD research tests, not yet discussed in detail, looked promising, but were not included in the final models. TAIL Distract, a component of a comprehensive auditory attention test for children (Zhang, Barry, Moore, & Amitay, 2012), showed a large significant influence on ASD prediction in this sample when paired with ECLiPS PSS and CCC SIDI (AUROC = 0.88).

However, the sample size was small (n = 29) and none of the predictor p-values was < 0.08 in the model. Similarly, the DLI-15 N, a measure from the Bergen Dichotic Listening Test (Moore et al., 2020), lifted the Attention AUROC to 0.97, when used with the SCAN Comp and CCC SIDI. But, again, the sample size was small (n = 32) and the p-value exceeded 0.1. Further validation of these findings could be sought in a larger-scale study.

LiD has been classified as an umbrella symptom of other assessments (e.g. APD, DLD, ADHD, ASD) rather than as a discrete disorder (Dillon & Cameron, 2021). Although LiD is a significant issue driving caregiver concerns, as shown by the ECLiPS, it was not specifically addressed by professions other than Audiology in this study. This disconnect, between presenting concerns and the specificity of assessment and intervention, may be a reason why APD originally emerged as a diagnostic category. Unfortunately, in the absence of specific and accepted assessment and treatment approaches, children with LiD can make the rounds to different clinical services and may receive help for language, attention or psychological factors that don’t address the complex verbal communication issues encountered in daily life at home and school.

Audiologists, due to their training and experience with hearing loss and device-oriented treatment approaches, are routinely sought out where there are concerns that a child may not be hearing well. Finding normal hearing function, audiologists have sought to explain listening problems that have no other clear explanation through challenging tests that stress the auditory system. While these tests document the extent of listening problems in some cases, they may also reflect non-auditory system problems that are more complex, multifactorial, and not easily addressed through audiologic management. Crucially, through longitudinal assessment, we have shown that LiD has long-lasting consequences, at least into adolescence (Kojima, 2024), and deserves to be addressed early and effectively with targeted treatment. Seeking input to this process through wider implementation of good quality caregiver reports could alert a variety of relevant clinical professionals to the needs of children with LiD.

## Conclusions

We show here that neither current research nor clinical test measures of LiD and APD are, considered separately, good predictors of clinical assessments and interventions to which they are currently directed. Caregiver report scores were more aligned with clinical practice. Some tests were predictive of clinical practice, but not in the discipline in which they are commonly used (e.g. the SCAN predicted a language diagnosis), and some test measures worked well in tandem with caregiver reports. Families of children with LiD have a wide range of concerns that extends well beyond the audiological domain. Audiologists and related professionals don’t know what to do about LiD since there is a dearth of clear evidence and agreed procedures, especially with respect to interventions. More research is needed to address these concerns and lack of knowledge, but caregiver reports provide inexpensive, simple and useful information to guide families and professionals towards appropriate outcomes.

## Supporting information

Supplemental Results

## Data Availability

All data produced in the present study are available upon reasonable request to the authors

## Acknowledgements

This research was supported by Grant R01 DC014078 (awarded to DRM) from the National Institute on Deafness and Other Communication Disorders and by the Cincinnati Children’s Research Foundation. DRM is also supported by the NIHR Manchester Biomedical Research Centre. The authors gratefully acknowledge the assistance of Audrey Perdew and Nicholette Sloat in testing children, and Susan Eichert, Sandra Grether, Diala Izhiman, Katie Effler, Alexandra Parshall, and Olivia Wnek in gathering, sorting and analyzing Epic data. The authors also thank the families who supported this project through participation as well as the Summer Undergraduate Research Foundation scholars who assisted with the project. We thank the two anonymous reviewers for their efforts in improving the quality of this paper.

## Footnotes

1 In this paper we use the term “condition” to refer to all clinical statements of impaired function since it was often unclear whether a formal diagnosis had been made. The exception is APD for which a clinical diagnosis was made in some cases. The term “assessment” is used to refer to research or clinical tests.

2 now known as “Sound Storm” (https://www.soundstorm.app/)

3 Four children who had previously been diagnosed with APD had total ECLiPS scores above this criterion (see Petley et al., 2021 for further discussion)

4 The Department of Developmental and Behavioral Pediatrics at CCHMC covers a range of disciplines of which only the Psychology Group is relevant here. We therefore refer to this team as “Psychology” through the rest of the study.

5 “Sensory processing” and “sensory processing disorder” are typically assessed by occupational therapists. They may involve any or all of the senses and can include up to 6 subtypes (STAR Institute). We analyzed the “Hearing and Auditory Processing” assessment with and without the 8 children who received this assessment.

6 All of the children in this study were found in the SICLiD study to have clinically normal hearing. However, 3 of them had been previously assessed with conductive hearing loss.

## Abbreviations

APD: Auditory Processing Disorder

AUROC: Area under the receiver operating characteristic

CCC-2: Children’s Communication Checklist (2nd edition)

CCC GCC: Children’s Communication Checklist – General Communication Composite

CCC SIDI: Children’s Communication Checklist – Social interaction difference index

CCHMC: Cincinnati Children’s Hospital Medical Center

DL: Bergen Dichotic Listening Test

DL l - 15: DL laterality score with an ILD = −15 dB

ECLiPS EAS: ECLiPS Environmental and Auditory Sensitivity subscale

ECLiPS: Everyday Children’s Listening and Processing Scale

ECLiPS LLL: ECLiPS Literacy/Language/Laterality subscale

ECLiPS MA: ECLiPS Memory and Attention subscale

ECLiPS PSS: ECLiPS Pragmatic and Social Skills subscale

ECLiPS SAP: ECLiPS Speech and Auditory Processing subscale

EMR: Electronic medical record

ILD: Interaural level difference

LiD: Listening Difficulties

LiSN-S: Listening in Spatialized Noise – Sentences test

LiSN HC: LiSN-S High Cue subscale

LiSN LC: LiSN-S Low Cue subscale

LiSN SA: LiSN-S Spatial Advantage subscale

LiSN TA: LiSN-S Talker Advantage subscale

NIH: National Institutes of Health Cognition Toolbox

NIH CC: Crystalized reasoning Composite score of NIH Cognition Toolbox

NIH DCCS: Directional Change Card Sorting test of NIH Cognition Toolbox

NIH ECC: Early Childhood Composite score of NIH Cognition Toolbox

NIH FC: Fluid reasoning Composite score of NIH Cognition Toolbox

NIH FT: Flanker test of NIH Cognition Toolbox

NIH LS: List sorting test of NIH Cognition Toolbox

NIH ORR: Oral reading recognition test of NIH Cognition Toolbox

NIH PC: Pattern comparison processing speed test of NIH Cognition Toolbox

NIH PV: Picture Vocabulary test of NIH Cognition Toolbox

NIH TC: Total Composite score of NIH Cognition Toolbox

OR: Odds ratio

PE: Tympanic pressure equalization tubes

SCAN AFG: SCAN Auditory Figure-Ground subscale

SCAN FW: SCAN Filtered Words subscale

SCAN CS: SCAN Competing Sentences subscale

SCAN CWD: SCAN Competing Words subscale

SICLiD: Sensitive Indicators of Childhood Listening Difficulties (study)

TD: Typically Developing

VIF: Variance inflation factor

## Notes

### Competing Interest Statement

David Moore is a paid consultant and stock holder of hearX Group

### Funding Statement

This study was funded by NIH DC014078, NIHR (UK), and the Cincinnati Children's Research Foundation

### Author Declarations

Ethics committee/IRB of Cincinnati Children's Hospital Medical Center gave ethical approval for this work

### Summary of Updates

All sections of the paper have been revised, except the Tables and Figures, and Supplemental Results have been added

