## Supplemental Results for "Multidisciplinary clinical assessment and interventions for childhood listening difficulty and auditory processing disorder: Relation between research findings and clinical practice"

JSLHR-24-00306
Multidisciplinary assessment and interventions for childhood auditory processing disorder (APD) and listening difficulties (LiD)

Supplementary Results

**Table S1.** Assessment codes allocated to electronic medical records (EMR; Epic) and the number of children receiving each code (Cases). All Hospital divisions.

|  | Cases |
| --- | --- |
| No clear assessment | 10 |
| APD, LiD | 19 |
| Attention, ADHD | 41 |
| Anxiety, panic | 15 |
| Social pragmatic | 5 |
| Language, speech, reading | 38 |
| Apraxia, dyspraxia | 2 |
| Developmental delay | 1 |
| Sensory processing | 7 |
| Adjustment disorder | 5 |
| Reading | 2 |
| Cognitive, intellectual | 5 |
| Academic | 1 |
| Hearing loss | 3 |
| Coordination, DCD | 6 |
| Syndrome | 1 |
| Impulsive, explosive | 1 |
| Oppositional defiant (ODD) | 4 |
| PTSD | 3 |
| Autism, ASD | 2 |
| Depression | 2 |
| Stuttering | 1 |
| Total | 164 |

**Table S2.** Summary assessments (from codes in Table S1)

|  | Cases |
| --- | --- |
| Language, speech, reading | 44 |
| Hearing, processing | 29 |
| Attention | 41 |
| Anxiety | 25 |
| ASD | 7 |
| Total | 146 |

**Table S3.** Speech/Language Pathology Intervention codes. Here, and in the following Tables S4-S6, the types of intervention in the EMR, either recommended or delivered, by each Hospital division, and the number of children receiving each code are listed.

|  | Cases |
| --- | --- |
| No record | 30 |
| No Intervention | 8 |
| APD | 3 |
| Speech sounds | 3 |
| Verbs | 1 |
| Pronouns | 1 |
| Multi-step directions | 9 |
| Sentences | 3 |
| Vocabulary | 2 |
| Intelligibility | 1 |
| Processing speed | 1 |
| Executive function | 1 |
| Language | 3 |
| Social skills, pragmatics | 6 |
| Tense | 2 |
| Articulation | 9 |
| Wh- yes/no questions | 6 |
| Phonemes | 3 |
| Word imitation | 1 |
| Labeling | 1 |
| Segment syllables | 1 |
| Word relations, stories | 3 |
| Sequence pictures | 1 |
| Word retrieval | 1 |
| Comprehension | 4 |
| Verbal expression | 1 |
| Figurative language | 1 |
| Location concepts | 1 |
| Reading | 6 |
| Writing | 2 |
| Error correction | 0 |
| Objects adjectives | 1 |
| Receptive/expressive | 5 |
| Recall exercises | 1 |
| Fluency | 2 |
| Auditory recall | 1 |
| Voice | 1 |
| Total | 96 |

Table S4 Audiology (see Table S3 for further details)

| Interventions | Cases |
| --- | --- |
| No record | 26 |
| FM, remote microphone | 5 |
| Accommodations | 6 |
| Computer training (Earobics) | 3 |
| Reading | 0 |
| Computer assisted instruction | 0 |
| No intervention | 25 |
| Memory training | 1 |
| Music training | 0 |
| Preteaching vocab | 1 |
| Dichotic listening training | 0 |
| SLT | 3 |
| Phonological awareness | 0 |
| Hearing loss | 1 |
| Compensatory skills | 0 |
| Communication strategies | 1 |
| Interhemispheric transfer | 1 |
| Auditory therapy | 0 |
| OT referral | 0 |
| ALD | 1 |
| psychoacoustic training | 0 |
| Total | 48 |

Table S5. Psychology (see Table S3 for further details)

| Intervention | Cases |
| --- | --- |
| No record | 56 |
| Medication (anxiety) | 2 |
| Medication (attention) | 4 |
| Medication (consider) | 1 |
| Classroom supports | 2 |
| Counseling | 2 |
| Family adjustments | 2 |
| CBT | 4 |
| Parent counseling | 2 |
| Behavior management | 3 |
| Care coordination | 1 |
| Listening | 1 |
| Impulse control | 3 |
| SLP | 1 |
| Education eval | 1 |
| Calming strategies | 2 |
| Environment mods | 1 |
| NOS GDD | 1 |
| Total | 33 |

Table S6. Occupational Therapy (see Table S3 for further details)

| Intervention | Cases |
| --- | --- |
| No record | 55 |
| Turn taking | 1 |
| Daily motor skills | 5 |
| Reading, comprehension | 1 |
| Self-help | 1 |
| Follow directions | 2 |
| Prompts, organization | 1 |
| Distraction, frustration | 3 |
| Preferential sitting | 1 |
| No specific therapy | 6 |
| Letters, writing | 2 |
| Spatial relations | 1 |
| Focus, attention | 1 |
| Soothing strategies | 2 |
| Family education | 1 |
| Total | 28 |

Table S7. Summary of interventions and the four most common interventions for each Division

| Division | Records | Interventions | % cases Intervened | Number diff. interventions | Most common intervention | 2nd most common | 3rd most common | 4th most common |
| --- | --- | --- | --- | --- | --- | --- | --- | --- |
| SLP | 44 | 36 | 81.8 | 35 | Multi step directions | Articulation | Pragmatics | Reading |
| Audiology | 48 | 23 | 47.9 | 19 | Accommodations | Computer training | Communication strategies | FM/remote microphone |
| Psychology | 18 | 17 | 94.4 | 16 | Medications (attention) | CBT | Behavior management | Impulse control |
| OT | 19 | 13 | 68.4 | 13 | Motor skills | Distraction | Follow directions | Writing letters |
| Total | 129 | 89 | 69.0 | 83 |  |  |  | |
